# Epigenetic biomarkers of ageing are predictive of mortality risk in a longitudinal clinical cohort of individuals diagnosed with oropharyngeal cancer

**DOI:** 10.1101/2020.02.04.20020198

**Authors:** Rhona A Beynon, Suzanne M Ingle, Ryan Langdon, Margaret May, Andy Ness, Richard Martin, Matthew Suderman, Kate Ingarfield, Riccardo Marioni, Daniel McCartney, Tim Waterboer, Michael Pawlita, Caroline Relton, George Davey Smith, Rebecca Richmond

## Abstract

**Background:** Epigenetic clocks are biomarkers of ageing derived from DNA methylation levels at a subset of CpG sites. The difference between predicted age from these clocks and chronological age (“epigenetic age acceleration”) has been shown to predict age-related disease and mortality. We aimed to assess the prognostic value of epigenetic age acceleration with all-cause mortality in a prospective clinical cohort of individuals with head and neck cancer: Head and Neck 5000.

**Methods:** We investigated two markers of intrinsic epigenetic age acceleration (*IEAAHorvath* and *IEAAHannum*), one marker of extrinsic epigenetic age acceleration (*EEAA*), one optimised to predict physiological dysregulation (*AgeAccelPheno*) and one optimised to predict lifespan (*AgeAccelGrim*). Cox regression models were first used to estimate adjusted hazard ratios (HR) and 95% confidence intervals (CI) for associations of epigenetic age acceleration with all-cause mortality in people with oropharyngeal cancer (n=408;105 deaths). The added prognostic value of epigenetic measures compared to a clinical model including age, gender, TNM stage and HPV status was then evaluated.

**Results:** *IEAAHannum* and *AgeAccelGrim* were associated with mortality risk after adjustment for clinical and lifestyle factors [HRs per standard deviation (SD) increase in age acceleration =1.32 (95% CI=1.08, 1.61; p=0.007) and 1.39 (95% CI =1.06, 1.83; p=0.017), respectively]. There was weak evidence that the addition of *AgeAccelGrim* to the clinical model improved 3-year mortality prediction (area under the receiver operating characteristic curve: 0.80 vs. 0.77; p-value for difference=0.069).

**Conclusion:** Our study demonstrates the potential of epigenetic age acceleration measures to enhance survival prediction in people with oropharyngeal cancer, beyond established prognostic factors.

## Introduction

Oropharyngeal cancer (OPC), which includes cancers of the soft palate, base of tongue, uvula, palatine tonsils and tonsillar pillars, is the second most commonly diagnosed head and neck cancer (HNC) in the UK after oral cavity cancer, with an overall age-standardised incidence rate of 2.9 per 100,000 persons ^1^. The main risk factors for OPC include smoking, alcohol consumption and human papillomavirus (HPV) infection ^2^. Estimated five-year survival rates for people with OPC vary from 35%-83% ^2 3^. As such, the ability to estimate survival probabilities at the time of diagnosis is important for clinical decision making and enrolment of low-risk individuals into treatment de-escalation trials, which aim to achieve similar efficacy as standard treatment regimens but with less toxicity and improved quality of life.

HPV positivity (primarily HPV16), is a major determinant of OPC prognosis ^4^. Compared to people with non-HPV-driven tumours, people with HPV-driven tumours have a 60% reduced risk of death 3-years post-diagnosis ^5^. Consequently, HPV status is now included in prognostic models alongside TNM stage and comorbidity. One such model has yielded a Harrell’s concordance statistic (C-statistic) of 0.68 (95% Confidence Interval [CI] 0.65, 0.71) in external validation, indicating good (but not excellent) prediction ^6^. The potential for model improvement is being explored and the prognostic value of lifestyle factors have been evaluated ^7-10^. The prognostic role of epigenetic biomarkers is less well studied.

Epigenetic biomarkers of ageing (“epigenetic clocks”), which are multivariate predictors of biological age based on DNA methylation (DNAm) levels at a subset of CpGs, are demonstrating promise in predicting age-related disease and mortality ^11 12^. Most studies evaluating the prognostic utility of these “epigenetic clocks” have been conducted in general (healthy) populations, however ^13 14^. There is a paucity of studies focusing on clinical populations. One study used a Cox model to estimate hazard ratios (HRs) for the association between epigenetic age acceleration (EAA), that is the difference between age predicted by the epigenetic clocks and chronological age, and risk of death following cancer diagnosis (n=1,726 deaths) ^15^. After adjusting for sociodemographic and lifestyle variables, the authors found limited evidence (OR=1.04, 95% CI: 1.00–1.09) of an association with EAA based on an epigenetic clock derived from methylation at 353 CpG sites (*EAAHorvath*) ^16^. However, mortality risk was 28% higher (OR=1.28, 95% CI: 1.11–1.47) for the highest versus lowest quartile of age acceleration based on an epigenetic clock derived from methylation at 71 CpG sites (*EAAHannum*) ^17^.

In this study, we aimed to investigate associations between all-cause mortality and epigenetic biomarkers of ageing in a clinical cohort of individuals diagnosed with OPC (n=408). In particular, we assessed associations between both “first generation” epigenetic clocks (*EAAHorvath* and *EAAHannum*) ^16 17^, derived from DNAm levels at CpG sites found to be strongly associated with chronological age, as well as more recently derived clocks: one optimised to predict physiological dysregulation (*AgeAccelPheno*) ^18^ and one optimised to predict lifespan (*AgeAccelGrim*) ^19^.

## Materials and methods

### Study population

The study population for this analysis included a subset of individuals enrolled in the Head and Neck 5000 (H&N5000) clinical cohort study, a UK-wide study of individuals with head and neck cancer (HNC). Full details of the study methods and overall population are described in detail elsewhere ^20 21^. Briefly, between April 2011 and December 2014, 5,518 individuals with HNC were recruited from 76 Head and Neck cancer centres across the UK. All people with a new diagnosis of HNC were eligible to join the study. Individuals with primary cancers of the pharynx, mouth, larynx, salivary glands and thyroid were included, while those with lymphoma, tumours of the skin or a recurrence of a previous HNC were excluded.

Consent was wide-ranging, including permission to: collect, store and use biological samples; carry out genetic analyses; collect information from hospital records and through self-reported questionnaires; and obtain mortality data through electronic record linkage. Ethics approval for the H&N5000 study was granted by the National Research Ethics Committee (South West Frenchay Ethics Committee, reference 10/H0107/57) and approved by the Research and Development departments of participating NHS Trusts.

Individuals were selected for analysis based on (pre-treatment) clinical coding of OPC (ICD-10 coding: C01, C05.1, C05.2, C05.8, C09.0, C09.1, C09.8, C09.9, C10.0, C10.2, C10.3, C10.8, C10.9), the availability of OncoChip genotype data, baseline questionnaire and data-capture information. Where possible, pathology reports of individual cases were subsequently checked to verify tumour site and subtype.

### Baseline data collection

Participants were asked to complete a series of three self-administered questionnaires at baseline enquiring about: 1) social and economic circumstances, overall health and lifestyle behaviours; 2) physical and psychological health, well-being and quality of life; and 3) past sexual history and behaviours. Information on diagnosis, treatment and co-morbidity was recorded on a short data capture form using questions based on a national audit ^22^. Clinical staging of the tumour was based on the American Head and Neck Society TNM staging criteria ^23^. In total, 5,474/5,518 (99%) data capture forms were completed and 3,361/5,385 (62%) individuals returned all three health and lifestyle questionnaires (133/5,518 participants enrolled in the study were either found to be ineligible or they did not consent/withdrew from completing baseline questionnaires [but remained in the study i.e. consented to data-linkage]).

Research nurses collected a blood sample from all consenting participants (n□=□4,676, 85%). Baseline sample collection was completed before the participants started their treatment, unless the individual’s diagnosis and treatment were one and the same, i.e. they were the same procedure (such as tonsillectomy). In this case recruitment and study baseline procedures were completed within a month of the diagnostic procedure. These were then sent to the study center laboratory (https://www.bristol.ac.uk/population-health-sciences/research/groups/bblabs/) at ambient temperature for processing. The blood samples were centrifuged at 3500 rpm for 10 minutes and the buffy coat layer used for DNA extraction. Any additional samples from the same participant were frozen and stored at - 80°C.

### Assessment of HPV status

The ‘gold standard’ test for oncogenic HPV infection is the expression of transcriptionally active HPV in fresh tissue ^24^. Tissue samples were not available for all H&N5000 participants in the current analysis. In the absence of available tumour specimens, the presence of circulating antibodies to type-specific HPV antigens appears to be a highly reliable diagnostic marker for HPV16-driven OPC with very high sensitivity and specificity ^25^. HPV serologic testing for HPV□16 (E6, E7, E1, E2, E4, and L1) antibodies was conducted at the German Cancer Research Center (DKFZ, Heidelberg, Germany) using glutathione S□transferase multiplex assays. HPV16 E6 seropositivity (a marker of HPV□transformed tumour cells ^26^), was indicated if HPV□16 E6 median fluorescence intensity (MFI) was >1000□units ^27^

### DNA methylation profiling

The participants were selected for DNAm profiling based on ICD-10 coding of OPC. To date, DNA samples isolated from buffy coats have been analysed for 448 participants. Following extraction, DNA was bisulphite-converted using the Zymo EZ DNA Methylation™ kit (Zymo, Irvine, CA, USA). Genome-wide methylation data were generated using the Infinium MethylationEPIC BeadChip (EPIC array) (Illumina, USA) according to the manufacturer protocol. The arrays were scanned using an Illumina iScan (version 2.3). Raw data files (IDAT files) were pre-processed using the R package *meffil* (https://github.com/perishky/meffil/) ^28^ to perform quality control (QC) and normalization, where control probes are utilised to separate biological variation from technical variation. Overall, 440/448 samples passed QC (sample exclusions: 2 samples with incorrect sex prediction, 3 samples with sex detection outliers, 1 sample with an outlier in predicted median methylated vs unmethylated signal, 2 duplicate samples) and were normalized. For the samples, the methylation level at each CpG site was calculated as a beta value (β), which is the ratio of the methylated probe intensity and the overall intensity and ranges from 0 (no cytosine methylation) to 1 (complete cytosine methylation).

### Estimation of epigenetic age

To generate the epigenetic ageing measures in H&N5000, we uploaded DNAm data for a subset of CpG sites from the Illumina EPIC array (n=27,523) to the online DNAm Age Calculator https://dnamage.genetics.ucla.edu/ developed by the Horvath laboratory. This subset of sites was chosen based on the list of 30,085 CpGs listed in the “datMiniAnnotation3.csv” file available for “Advanced Analysis” on the DNAm Age Calculator website. 2,562 CpG sites were missing due to probe discrepancy between the Illumina EPIC platform and Illumina 450K platform, the latter of which was used to derive some of the epigenetic clocks. We also uploaded an annotation file, containing data on chronological age, sex and tissue type for the samples. We were able to generate the following epigenetic ageing measures for 440 individuals (following the notation of previous publications): intrinsic epigenetic age acceleration based on Horvath’s multi-tissue predictor (*IEAA*) ^16^; intrinsic epigenetic age acceleration based on Hannum’s predictor (*IEAAHannum*) ^17^; extrinsic epigenetic age acceleration (*EEAA*), an enhanced version based on Hannum’s method, which up-weights the contribution of blood cell composition ^12^; PhenoAge (*AgeAccelPheno*) ^18^ and GrimAge (*AgeAccelGrim*) ^19^. *AgeAccelPheno* and *AgeAccelGrim* can be considered as measure of extrinsic aging on the basis of estimators used ^19 29^. An overview of the different epigenetic age predictors is provided in Table 1. Intrinsic epigenetic age acceleration (IEAA) is independent of changes in blood cell composition while extrinsic epigenetic age acceleration (EEAA) incorporates age-related changes in blood cell composition ^12^.

**Table 1:** Overview of various measures of epigenetic age acceleration.

| Measure of epigenetic age acceleration | Abbreviation | CpGs | Description |
| --- | --- | --- | --- |
| Intrinsic Epigenetic age acceleration based on Horvath | *IEAA* | 353 | The residual resulting from regressing DNAm age on chronological age and estimates of major blood immune cell counts<sup>*</sup>. |
| Intrinsic epigenetic age acceleration based on Hannum | *IEAA Hannum* | 71 |  |
| Extrinsic age acceleration based on Hannum | *EEAA* | 71 | The residual resulting from a univariate model regressing a weighted age estimate (which increases the contribution of 3 cell types known to change with age<sup>**</sup>) on chronological age. |
| Age acceleration based on AgePheno | *AgeAccelPheno* | 513 | The residual resulting from a linear model when regressing *PhenoAgeAccel* on chronological age, where *PhenoAge* is an ageing measure based on a linear combination of chronological age and nine clinical biomarkers. |
| Age acceleration based on AgeGrim | *AgeAccelGrim* | 1,030 | The residual resulting from a linear model when regressing *AgeGrim* on chronological age, where *AgeGrim* is an ageing measure based on a linear combination of chronological age, gender, and DNAm-based surrogate biomarkers for smoking pack-years (*dnampackyears*) and seven plasma protein levels. |

*\* Naive CD8+ T cells, exhausted CD8+ T cells, plasmablasts, CD4+ T cells, natural killer cells, monocytes, and granulocytes.*
*\*\* naïve (CD45RA+CCR7+) cytotoxic T cells, exhausted (CD28-CD45RA-) cytotoxic T cells, and plasmablasts.*

### Study follow-up and survival

Regular vital status updates were received from the NHS Central Register (NHSCR) and NHS Digital (formally called the Health and Social Care Information Centre (HSCIC)), notifying on subsequent cancer registrations and death among cohort members in the H&N5000. Recruitment for the study finished in December 2014 and follow-up information on survival status was obtained on 1^st^ September 2018. The median duration of follow-up was 4.3 years (Interquartile range (IQR): 3.3 to 5.2).

### Covariates

Information on baseline age at consent into the cohort, sex, weight, height, highest educational attainment (School education, college or degree-level), smoking status and alcohol intake were obtained from self-completed questionnaires, which are available on the study website (http://www.headandneck5000.org.uk/). Body mass index (BMI) was calculated based on participants’ self-reported height and weight, using the equation: weight (kg) / [height (m)]^2^. Age and BMI were treated as continuous variables. Smoking status was defined as “current”, “former” or “never” user of tobacco, whereby the definition of a never-user was someone who had never consumed at least one tobacco product per day for a year. Participants were asked to report the number of alcoholic drinks they consumed on an average week before they became ill, providing information separately for wine (in glasses), spirits (in measures), and beer/larger/cider (in pints). Initially, drinks were converted into grams of ethanol. The sum of these converted measures was then used to calculate the number of units of alcohol consumed in a typical week, where 1 UK unit translates to 10ml, or 8g, of pure alcohol. Baseline alcohol drinking categories (males and female) were then defined as none, moderate (≤14 units/week), and hazardous to harmful (>14 units/week), based on current UK guidelines.

Information on ICD code, TNM staging and comorbidity were extracted from the baseline data capture forms. Comorbidity was described using the Adult Comorbidity Evaluation-27 (ACE-27), a validated comorbidity instrument that provides a score (0–3) based on the number and severity of medical comorbidities ^30^. For analysis, a categorical comorbidity variable was derived corresponding to extent of decompensation (functional deterioration)-“none”, “mild”, “moderate” or “severe”.

It was decided *a priori* not to include ethnicity as a covariate in the analysis because only 2 individuals reported being white.

### Statistical analysis

Stata (Release 15.1, StataCorp) was used for all analyses reported below. The analysis was split into two parts. Firstly, we examined whether the EAA measures were associated with survival, after controlling for established HNC prognostic factors (listed below); secondly, we investigated whether these EAA measures provide any additional prognostic information, over and above those factors that are currently considered routinely in clinical practice, namely age, gender, tumour stage, HPV status and comorbidity. All tests of statistical significance should be two sided

#### Step 1: examining the association of EAA measures with survival

Descriptive analyses were first performed to explore the distribution of, and correlations between EAA measures, using histograms and Pearson’s correlation coefficients. Baseline descriptive data were stratified by survival status at three years. The univariate association of covariates on all-cause mortality risk was assessed using Kaplan-Meier curves and the log-rank test.

Multivariable Cox proportional hazards models were then used to examine the potential associations of our epigenetic age measures with overall survival, defined as the time in years from study enrolment to date of death from any cause or date of censorship (i.e., the last date of follow-up). Cancer-specific mortality data were not available at the time of data analysis.

Given that the five epigenetic measures of age acceleration were expressed in different units, we standardised measures using z-scores to allow comparison of effect estimates. (HRs) and 95% CIs for all-cause mortality were calculated for each standard deviation (SD) increase in epigenetic age acceleration.

For each measure of epigenetic age acceleration, four separate Cox models were run: 1) a minimally adjusted model that only controlled for gender; 2) a model that additionally controlled for clinical factors (TNM stage, HPV status, comorbidity and BMI); 3) a model that additionally controlled for socio-demographic and economic factors (education, annual household income, marital status) and 4) a fully adjusted model that additionally controlled for modifiable lifestyle behaviours (smoking and alcohol consumption). Models were selected *a priori* based on the existing literature linking these covariates with survival ^8 31-33^

Several of our covariates of interest had some missing data, particularly BMI as this measure was not initially collected at recruitment into the H&N5000 study. Excluding individuals with missing covariate data would have reduced statistical power to detect an association between the epigenetic age measures and survival, and so multiple imputation (MI) was performed. Previous work suggests that MI provides unbiased results in situations where data are missing at random (MAR) ^34^, i.e., any systematic differences between the observed and missing data can be explained by associations with the observed data. Missing values were imputed using the ICE package for multiple chained equations in Stata ^35^. Twenty imputed datasets were generated and analysed separately using standard statistical methods and the multiple sets of results combined using ‘Rubin’s rules’ ^36^. The imputation models contained all the variables in the analysis model (including the outcome) and the Nelson–Aalen estimator of the cumulative hazard. As a sensitivity analysis, we conducted a complete case analysis including only those participants with data available for our covariates of interest, and analysed as above ^37^.

#### Step 2: assessing the prognostic value of EAA measures

Evidence of an association with survival is not enough to include novel biomarkers in prediction models; to be useful to clinicians they must provide added prognostic value to existing models. We therefore explored whether the addition of EAA measures to existing models based on established mortality risk factors (i.e. those currently considered in clinical decision making), improved model performance.

Survival models were fitted using the methods of Royston and Parmar ^38^which model the baseline hazard (on the log-cumulative hazard scale) using restricted cubic splines. These are known as flexible parametric survival models. Unlike Cox models, these models permit absolute (as opposed to relative) measures of effect (i.e. survival probability) to be estimated at all time points, rather than just at event times and thus it is easy to obtain predictions, both in and out of sample. It also allows time-dependent effects to be modelled. The Royston and Parmar models were fitted using maximum likelihood estimation via the ‘stpm2’ command in Stata. The spline complexity for the baseline hazard which best fits the data was investigated visually and through model fit statistics [Akaike Information Criterion (AIC) and Bayesian Information Criterion (BIC)]. We considered possible degrees of freedom (df) ranging from 1□to 5□df (for a model with no variables included). Using the hazard function plots and the AIC and BIC as a guide, 2 df were deemed sufficient. These 2 degrees of freedom equate to 1 interior knot in the baseline hazard. Non-linear relationships with continuous predictors were considered using the multivariable fractional polynomial (MFP) algorithm described by Sauerbrei and Royston ^39^ and implemented in Stata using the ‘mfp’ command.

The following models were fit: 1) a ‘clinical model’, which comprised age, gender, TNM stage, HPV status and comorbidity; 2) clinical + *IEAA*; 3) clinical + *EEAA*; 4) clinical + *IEAAHannum*; 5) clinical + *AgeAccelGrim* and 6) clinical + *AgeAccelPheno*. Models were fit in a sub-sample of participants with complete data available for the clinical covariates considered in this analysis (age, gender, tumour stage, comorbidity and HPV status). The performance measures examined were the AIC, which measures the relative goodness of fit of a model, considering both the statistical goodness of fit and the number of parameters used, and the Harrell’s concordance statistics (or C-statistics), an extension of the area under the receiver operating curve (AUC) to survival analysis ^40 41^. The interpretation of the statistic is equivalent, namely, a C-statistic of 0.5 indicates no discrimination above chance (of dying or surviving), whereas a C-statistic of 1.0 indicates perfect discrimination, and thus superior prediction.

ROC curves and AUC functions were calculated to characterize how well the models distinguished between participants for whom the event (death) did and did not occur three years after diagnosis. Internal validation was performed on the final model using 500 bootstrap samples to adjust performance for optimism and calculate a shrinkage factor to be applied to the models’ regression coefficients for use in other (external) settings.

## Results

Of the 1,896 H&N5000 participants with OPC, 408 had epigenetic data available (32/440 available for analysis were excluded based on pathological re-coding, i.e. cancers did not originate in the oropharynx; Figure 1). A total of 105 deaths were observed during follow-up [median=5.3 years, interquartile range (IQR) 4.9 to 6.0].

**Figure 1:**
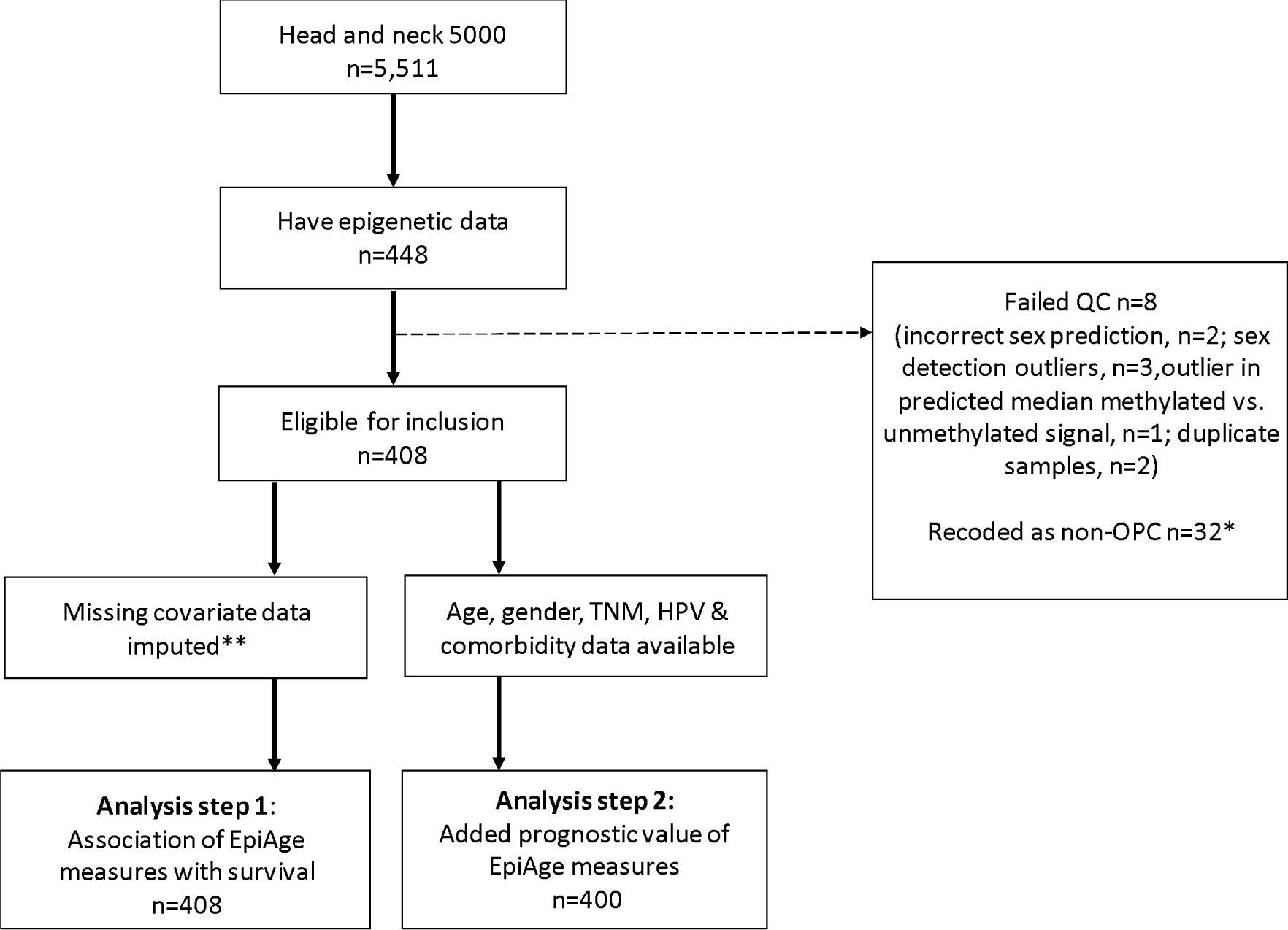
flow of participants included in the analysis. Based on pathological ICD coding.

### Missing data

The proportion of missing data is summarised in Supplementary Table 1. The largest proportion of missing data corresponded to BMI (33.3%), followed by annual household income (12.5%) and education (4.7%).

### Baseline descriptives

Table 2 shows the baseline characteristics of participants. There were differences in age (*p*-value for difference < 0.001), BMI (*p*=0.001), HPV status (*p*=<0.001), comorbidities (p=0.001), smoking status (*p*=<0.001), alcohol consumption *(p*=0.019), annual household income (*p*=0.006) and marital status (*p*<0.001) between participants who were and were not alive three years after enrolment into the study. Participants who were alive at three years had a mean age of 57.4 years at the time of entry (standard deviation (SD) = 8.9) compared to 62.9 years (SD = 11.3) for those who were dead at three-years. Mean epigenetic age acceleration, as measured by each of the epigenetic clocks, was higher in those individuals who had died at three years. There were differences between people who had survived and those who had died for the following epigenetic age measures: *EEAA* (*p*=0.004), *IEAAHannum* (*p*=0.006), *AgeAccelPheno* (*p*=<0.001) and *AgeAccelGrim* (*p*=0.002). Corresponding results for participants included in the complete case analysis can be found in Supplementary Table 2.

**Table 2:**
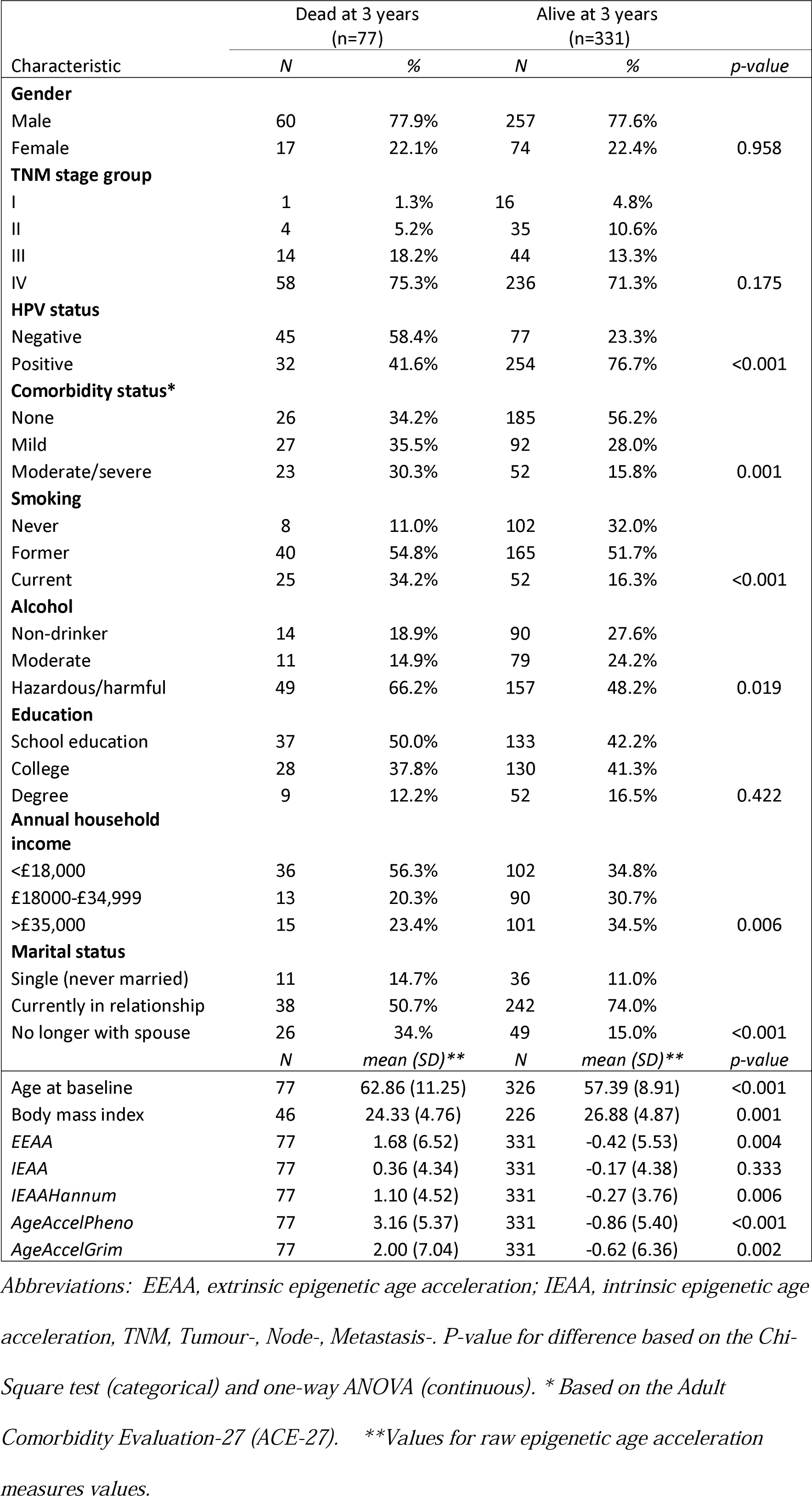
Baseline characteristics of the study sample stratified by 3-year mortality status.

### Pairwise correlations between measures of epigenetic age acceleration

The pairwise Pearson correlation coefficients of selected measures of *EAA* is shown in Supplementary Figure 1. The measures exhibited correlations ranging from 0.05 to 0.74. the strongest absolute association was between *EEAA* and *IEAAHannum* with the majority of correlations ranging between 0.33 and 0.54.

### Association of DNA Methylation-based biological age with survival

The results of the minimally adjusted and fully adjusted Cox regression analyses on imputed data (n=408) are presented in Figure 2. An overview of all the model outputs is provided in the supplementary material (Supplementary Tables 3 and 4 for the imputed and complete case analysis, respectively).

**Figure 2:**
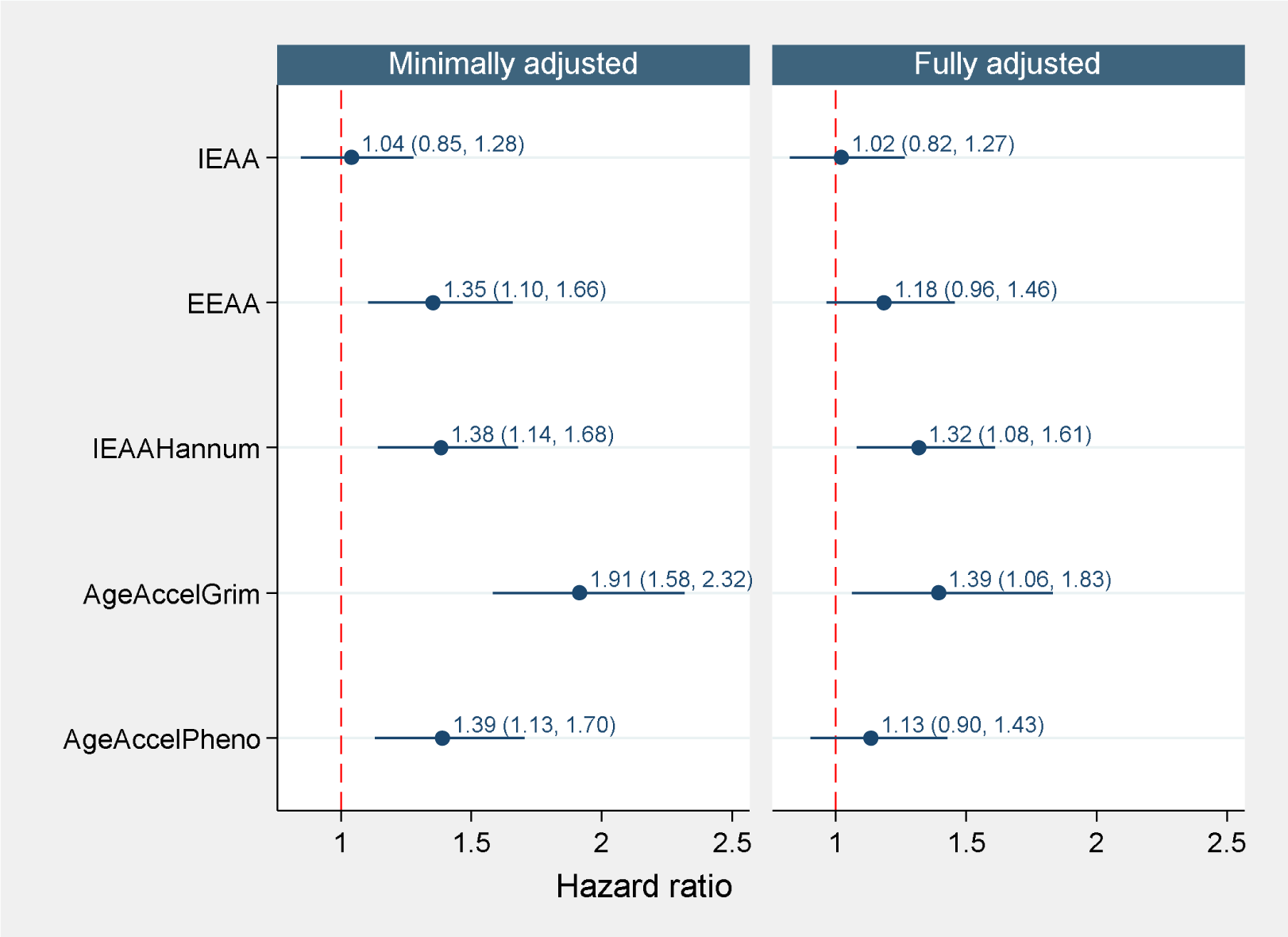
Association of epigenetic age acceleration measures with mortality risk (n=408). Minimally ajusted model controlled for sex. Fully adjusted model additionally controlled for TNM stage, HPV status, comorbidity, BMI, educational attainment, annual household income, marital status, smoking, and alcohol intake.

In the basic model, which adjusted only for sex, there was evidence that all of the epigenetic biomarkers of aging were associated with survival, except for one intrinsic epigenetic age acceleration measure (*IEAA*). All of the reported associations are in the expected directions, e.g. higher values of EAA are associated with higher mortality risk. HRs ranged from 1.35 (95% CI=1.10, 1.66; *p*=3.5 ×10^−03^) for *EEAA* to 1.91 (95% CI=1.58, 2.32; *p*=2.3 ×10^−11^) for *AgeAccelGrim* (Figure 2), where HRs represent the difference in mortality risk per SD unit increase in EAA measure. For *IEAAHannum* and *AgeAccelGrim*, evidence of associations remained but were attenuated following adjustment for clinical and socioeconomic factors (Figure 2). In the fully adjusted model, which also adjusted for smoking and alcohol intake, *IEAAHannum* and *AgeAccelGrim* were still associated with mortality risk (HRs=1.32 (95% CI=1.08, 1.61; *p*=6.9 ×10^−03^) and 1.39 (95% CI=1.06, 1.83; p=0.017), respectively (Figure 2).

In the complete case analysis (n=225; 49 deaths), the results of the basic model were broadly comparable to those of the imputed analysis (Supplementary Table 4). However, *IEAAHannum* was not robust to adjustment for socioeconomic factors and the association of *AgeAccelGrim* with survival attenuated following adjustment for smoking and alcohol intake.

### Examination of the predictive utility of EAA measures at 3-years

Table 3 shows the performance measures for the fitted models. The AIC values for the clinical + *EAA*, clinical + *IEAAHannum* and clinical + *AgeAccelGrim* models were lower than that of the standard clinical model which does not include measures of EAA. As a rule of thumb, two models are generally considered equivalent if the difference in their AICs is less than 2 units ^42^; therefore on this basis, all three of these models had a better overall fit compared to the standard clinical model. C-statistics ranged from 0.75 for the clinical model to 0.80 for the clinical + *AgeAccelGrim* model.

**Table 3:** Measures of model performance.

| Model | AIC | C-statistic (95% CI) |
| --- | --- | --- |
| Clinical | 486.93 | 0.75 (0.70, 0.80) |
| Clinical + *EEAA* | 483.36 | 0.76 (0.71, 0.81) |
| Clinical + *IEAA* | 488.14 | 0.76 (0.71, 0.81) |
| Clinical + *IEAAHannum* | 480.10 | 0.77 (0.72, 0.82) |
| Clinical + *AgeAccelGrim* | 473.14 | 0.78 (0.73, 0.83) |
| Clinical + *AgeAccelPheno* | 485.52 | 0.76 (0.71, 0.81) |

*Abbreviations: AgeAccelGrim, age acceleration based on DNAmGrimAge; AgeAccelPheno; age acceleration based on PhenoAge; AIC, Akaike's information criterion; C-statistic, Harrel's concordance statistic; EEAA, extrinsic epigenetic age acceleration; IEAA, intrinsic epigenetic age acceleration; 95% CI, 95% confidence interval*

Given that the clinical + *AgeAccelGrim model* showed the strongest association in the Cox regression analysis, appeared to fit the data best and yielded the highest discrimination (i.e. had the lowest AIC and highest C-statistic), we examined whether this model provided improved prediction at three years compared to a clinical model (including age, sex, TNM stage, HPV and comorbidity), by comparing AUC values. The results are presented in Figure 3. There was weak evidence to suggest the clinical + *AgeAccelGrim* model had superior predictive performance compared to the clinical model (clinical AUC: 0.77, clinical + *AgeAccelGrim* AUC: 0.80; p-value for difference=0.069), at three years after diagnosis when there had been 76 deaths. The bootstrap optimism corrected AUC values showed a small reduction in performance compared with the original model (optimism adjusted AUCs of 0.74 and 0.77 for clinical and clinical *+ AgeAccelGrim* models, respectively).

**Figure 3:**
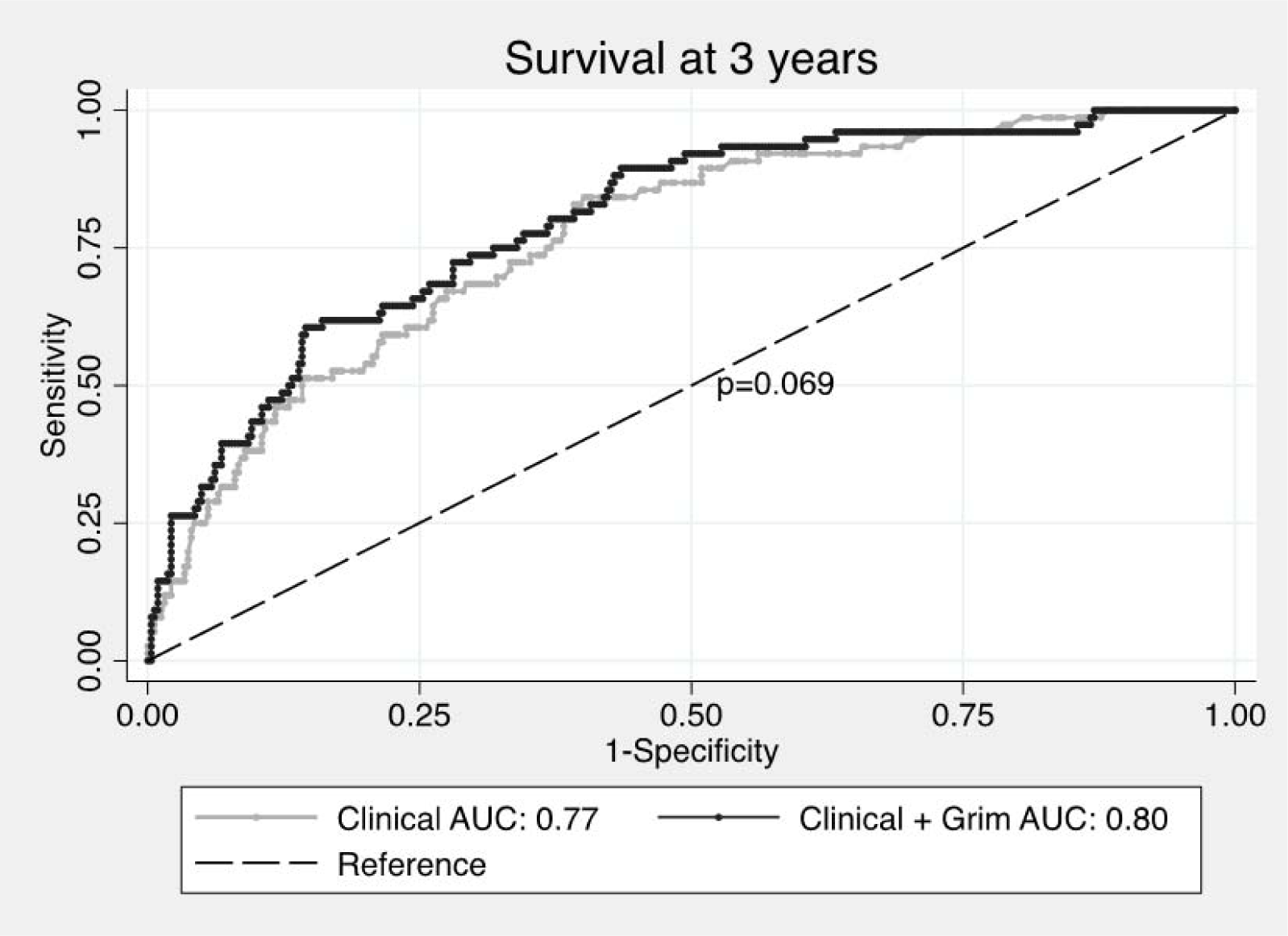
Independent contribution of AgeAccelGrim to prognosis beyond clinical factors. Abbreviation: AUC, area under the receiver operating curve statistic.

The optimism-adjusted c-slope (or uniform shrinkage factor) for the *clinical + AgeAccelGrim* model, was 0.83, indicating there was some overfitting. The original predictor effects (beta coefficients) were adjusted by this value ^43^. The results are presented in Table 4. In the adjusted model, each SD unit increase in *AgeAccelGrim* was associated with a 1.5-fold increased risk of death at 3 years (optimism adjusted HR: 1.54, 95% CI=1.2, 1.92; p=<0.001).

**Table 4:** Estimated coefficients (uncorrected and corrected) for the clinical + AgeAccelGrim model.

| Variable | Original model |  |  | Final model after adjustment for overfitting |  |  |
| --- | --- | --- | --- | --- | --- | --- |
|  |  | 95% CI |  |  | 95% CI |  |
| | $\beta$ | ll | ul | $\beta$ | ll | ul |
| **Age** | 0.05 | 0.02 | 0.07 | 0.04 | 0.02 | 0.06 |
| **Gender** |  |  |  |  |  |  |
| Female | 0.42 | -0.14 | 0.99 | 0.35 | -0.12 | 0.82 |
| **Tumour stage** |  |  |  |  |  |  |
| II | 0.64 | -1.56 | 2.85 | 0.53 | -1.29 | 2.36 |
| III | 1.65 | -0.38 | 3.69 | 1.37 | -0.32 | 3.06 |
| IV | 1.85 | -0.14 | 3.84 | 1.54 | -0.12 | 3.19 |
| **HPV status** |  |  |  |  |  |  |
| Positive | -0.95 | -1.47 | -0.44 | -0.79 | -1.22 | -0.36 |
| **Comorbidity\*** |  |  |  |  |  |  |
| mild | 0.33 | -0.23 | 0.90 | 0.28 | -0.19 | 0.75 |
| moderate/severe | 0.24 | -0.38 | 0.85 | 0.20 | -0.31 | 0.70 |
| **AgeAccelGrim** | 0.52 | 0.26 | 0.78 | 0.43 | 0.22 | 0.65 |

Regression coefficients ( $\beta$ ) and 95% confidence intervals (CI) for 3-year overall survival.

Smoking has been shown to be independently predictive of mortality in Head and Neck 5000 ^9^. The reduced effect estimate observed between *AgeAccelGrim* and mortality with adjustment for smoking status suggests that the enhanced prognostic ability gained from adding *AgeAccelGrim* to the clinical model could be due to the inclusion of a smoking predictor (Table 1) ^19^. In order to investigate this, we conducted an additional sensitivity analysis whereby we compared the prognostic ability of the following models: 1) clinical + *AgeAccelGrim*; 2) clinical + self-reported smoking; and 3) clinical + *dnampackyears*, the DNAm-based surrogate biomarker for pack years of smoking used to derive AgeGrim (n=384 participants with smoking data available; no. deaths=72). At 3-years, there was a suggestion that the clinical + *AgeAccelGrim* model had better discrimination (AUC value of 0.80) than the clinical models including both self-reported smoking (AUC=0.77) and a DNAm surrogate for pack years (AUC=0.78), although there was limited evidence of a difference in AUCs based on chi-squared tests (p=0.175) (Supplementary Figure 2).

## Discussion

In this study of 408 oropharyngeal cancer cases with a median of 5 years of follow-up, we demonstrate that two different epigenetic age estimators, *IEAAHannum* and *AgeAccelGrim*, are associated with increased risk of all-cause mortality and these associations are independent of known mortality risk factors, including several clinical, demographic and lifestyle variables. *AgeAccelGrim* had the strongest effect estimate, with each SD increase of epigenetic age acceleration (relative to chronological age) resulting in a 39% increase in risk of death in the fully adjusted model (HR=1.39; 95% CI: 1.06, 1.83). When compared to a standard clinical model that included age, sex, tumour stage, HPV status and comorbidity, the addition of *AgeAccelGrim* to the model showed some improvement in mortality risk prediction at 3-years (clinical AUC: 0.77, clinical + *AgeAccelGrim* AUC: 0.80; *p*=0.069).

Survival for people with OPC is heterogeneous. As such, there is continued interest in identifying new prognostic factors (clinical, lifestyle, genetic, and epigenetic) that can improve our ability to predict disease and mortality outcomes ^44 45^. Among healthy populations, DNA methylation-based measures of epigenetic aging have shown great promise for predicting mortality ^11 12^ and yet, whilst the accumulation of epigenetic changes is a hallmark of cancer, few studies have prospectively examined the potential of epigenetic clocks to predict mortality among people with cancer, including HNC.

Our findings are in line with the current literature, which suggests that DNA-methylation derived “GrimAge”, a composite biomarker incorporating DNA methylation - based surrogates for smoking pack-years and seven plasma proteins as well as age and sex, is a better predictor of mortality risk compared to first-generation DNA methylation-based predictors (i.e. the epigenetic clocks developed by Horvath and Hannum) ^19^. Age acceleration as measured by GrimAge not only performs better in predicting time to death, time to coronary heart disease and time to cancer among general populations, but also shows associations with established risk factors ^19^.

The finding that *AgeAccelGrim* was most strongly related to mortality risk in the current study may have been in part related to the inclusion of the surrogate measure for smoking in the GrimAge biomarker. This is because smoking has been shown to be independently predictive of mortality among HNC cases ^9^. When we compared the prognostic performance of the clinical + *AgeAccelGrim* model with clinical models including both self-reported smoking and the DNAm surrogate biomarker for pack years of smoking, the clinical + *AgeAccelGrim* model had better discrimination. These findings suggest firstly, that the methylation-based measure of smoking is a better indicator, with less misclassification, than self-report, and secondly, that the prognostic utility of *AgeAccelGrim* does not appear to be solely driven by the inclusion of the DNAm-based biomarker for smoking, although the improvement in predictive performance was only marginal.

In addition to the inclusion of a surrogate measure for smoking, GrimAge is trained on a small set of proteins known to be associated with mortality ^19^, including those of plasminogen activator inhibitor 1 (PAI-1) and growth differentiation factor 15 (GDF15). PAI-1 (aka SERPIN E1) is overexpressed in a variety of tumours and has been found to be a strong predictor of poor clinical outcome and poor response to ^46^, whilst GDF15 is involved in the pathogenesis of oral squamous cell carcinoma (OSCC) ^47^.

Our investigation has several strengths including the relatively long follow-up period, the fact that individuals were sampled at the time of diagnosis and the fact that DNA methylation of the samples was assayed simultaneously in the same laboratory, thus minimising potential technical bias. We were also able to account for a range of factors that could confound our effect estimates, including smoking, alcohol intake and BMI, all of which are known to influence DNAm and HNC risk. Moreover, missing covariate data were imputed via chained equations to minimise possible biases.

Although we found evidence that *IEAAHannum* and *AgeAccelGrim* were associated with higher mortality risk, our study has several limitations. First, the sample size for our analysis was relatively small and we were unable to identify independent prospective datasets to validate our findings. In smaller datasets, prediction statistics are more easily influenced by a small number of observations, and consequently, the results observed here could be due to chance. In order to mitigate this, we obtained estimates of a uniform shrinkage factor (the average calibration slope) and multiplied this by the original β coefficients from the fitted model to obtain optimism adjusted coefficients. Second, it was not possible to examine cancer-specific mortality since cause of death data are not currently available for all participants. Third, information on some of the variables used in our analysis, were obtained via participants’ self-report (e.g. BMI, educational attainment, smoking and alcohol intake), which can result in recall bias or misreporting. We utilised a DNAm-derived measure of packyears of smoking in our sensitivity analysis but future studies could implement the use of other methylation scores to index these variables ^48 49^. Fourth, there is a disparity in coverage between 450K and 850K Illumina platforms meaning that some of the CpGs included in Horvath’s and Hannum’s clock are missing. This could be problematic, although a previous study examining the application of EPIC array data to predict DNAm age demonstrated that the lack of the clock-CpGs on the EPIC array did not undermine the utility of the epigenetic age predictors ^50^. Finally, we did not account for multiple testing of the 5 epigenetic age acceleration measures, although evidence of correlation between some of the epigenetic measures suggests that correction for multiple independent tests may not have been appropriate.

## Conclusion

Overall, our findings provide evidence that DNA methylation-based estimators of ageing could provide prognostic utility, above established prognostic factors including age, sex, tumour stage, HPV status, comorbidity and smoking. That an accurate and easy-to-measure biomarker derived from peripheral blood could serve as a better predictor of mortality risk in people diagnosed with OPC is important as this could impact treatment planning and provide information that improves patient stratification in study design, e.g. treatment de-escalation trials. Nonetheless, these findings should be further investigated in a larger, independent sample and including other ethnicities.

## Data Availability

Head and Neck 5000 is run as a resource to be used by the research community. There is a policy for access to the resource, which can be found on the study website: http://www.headandneck5000.org.uk/information-for-researchers/usingtheresource/

## Abbreviations

AIC: Akaike’s information criterion
AUC: Area under the receiver operating characteristic curve
BIC: Bayesian Information Criterion
CI: Confidence interval
CpG: Cytosine-phosphate-Guanine site
EAA: epigenetic age acceleration
HNC: Head and neck cancer
HPV: Human papillomavirus
HR: Hazard ratio
IQR: Inter-quartile range
MAR: Missing at random
OPC: Oropharyngeal cancer
SD: Standard deviation

## Acknowledgements

The authors are extremely grateful to all Head and neck 5000 participants, the Head and Neck 5000 study co□ordination team and the laboratory technicians at the Bristol Bioresource Laboratories.

